# SOUTH AMERICAN INDIGENOUS COMMUNITIES AND BODY MASS INDEX: A SYSTEMATIC REVIEW AND META-ANALYSIS

**DOI:** 10.1101/2021.02.04.21251146

**Authors:** Carlos P Jara, Natalia Ferreira Mendes, Claudinei José Gomes Campos, Maria Isabel Pedreira de Freitas, Henrique Ceretta Oliveira, Lício Augusto Velloso, Eliana Pereira de Araujo

## Abstract

Obesity is an increasing new pandemic. Currently more than 1.9 billion adults are overweight and at least 700 million of them are obese. Obesity is the result of a positive energy balance, which is conditioned by both environmental and genetic factors. Interestingly, individuals from similar ethnic-based ancestry communities, share both environmental and genetic features. Here, we described the relationship between indigenous Chilean groups and body mass Index. We conducted a Systematic review and Meta-analysis on Pubmed, LILACS, Scielo, Web of Science and Scopus databases. Our results showed that Indigenous Children present a lower BMI than Non-Indigenous Children. However, no difference within BMI was identified in adults. The gender affected the BMI as well. Aymara and Mapuche Women presented higher BMI than Indigenous Men. In the other hand, Indigenous people living in rural environment showed lower BMI than those whose live-in urban areas. Finally, Indigenous communities presented no difference in the risk to suffer Obesity when compared with Non-Indigenous communities. Here suggest that ethnicity could be a health determinant as well as a risk factor for obesity. Then, targeted prevention strategies with ethnic-based focus would be developed.

## Introduction

Obesity is a new pandemic of the 21st century. Currently, more than 1.9 billion adults are overweight and at least 700 million of them are obese (1, 2). In Chile, general population has progressively increased the average of Body Mass Index (BMI) during the last four decades going from normal to overweight (3). The main challenge related to obesity is the risk to suffer Cardiovascular Diseases (CVDs) (2). CVDs are the leading cause of general mortality and disability worldwide (1).

Obesity is the result of a positive energy balance, determined at least by both environmental and genetic factors. Interestingly, individuals within ethnic-based ancestry communities, share both environmental and genetic features (4-8). Specifically, previous reports have explored the relationship between ethnicity and obesity (9-12). For instance, it was showed the relationship between Mapuche ancestry, a native of South America ethnicity, presented the lowest prevalence of Type 2 Diabetes (T2DM) with 1.4%. In the other hand, a North American indigenous group, The Pima community, showed the highest T2DM prevalence reaching 50% of them (13). Mapuche indigenous group represent the vast majority of all the Chilean Indigenous population with 87% of the total indigenous population followed by Aymara indigenous community (4). These communities are highly concentrated in the north (Aymara), and the south (Mapuche) of Chile (7). Until now, there are more than 20 years of cross-sectional BMI reports of Mapuche and Aymara communities. However, these reports have been inconclusive and sometimes contradictories. For this reason, our objective was to identify the relationship between Mapuche and Aymara groups and BMI and obesity.

## Methodology

Systematic review and Meta-analysis conducted in Pubmed, LILACS, Scielo, Web of Science and Scopus databases using a mixed strategy-key words: mapuche OR aymara AND (BMI OR “Body Mass Index” OR obesity OR weight). Two authors, CPJ and NFM, independently identified cross-sectional and longitudinal studies published before February 02, 2021, that reported BMI. We used the World Health Organization criteria of obesity to adults: BMI ≥ 30.

### Criteria for inclusion

Original articles in English with primary source of BMI expressed in Mean and Standard Deviation (SD) or Confidence Interval (CI). Indigenous group criteria (Aymara or Mapuche) were in according to the Article 2 and 3 of the 19,253 Chilean Indigenous Law (November 2017): 1) Those who are sons of an indigenous father or mother 2) if their last names (father’s surname and/or mother’s maiden name) were of Mapuche or Aymara origin. 3) Those who maintain cultural features of Aymara or Mapuche ethnic groups, understanding as such the practice of lifestyle, customs, or religion of these ethnic groups in a habitual way or whose spouse is indigenous. In these cases, it will be necessary, in addition that they self-identify as indigenous. Mapuche indigenous group included Pehuenches, Picunches, Huilliches and Lafquenches groups. Non-indigenous criteria were determined as Caucasian and European origin. Aymara and Mapuche surnames are recognizably different from European ones. Even more, Chilean indigenous surnames have been validated as sufficient information to determine indigenous genetic background (14). The geographical region included was South America. Age: 1 year to 100 years. Gender: social/self-recognized as a man, social/self-recognized as a woman, social/self-recognized as a transgender or social/self-recognized as a genderless.

### Criteria for exclusion

Pregnancy, Type 1-2 Diabetes Mellitus, Control group mixed with ethnics group and people with unhealthy status.

### Level of evidence

It was determined by the National Health and Medical Research Council (NHMRC) Evidence Hierarchy (15). The Flow Diagram was based on the *Preferred reporting items for systematic reviews and meta-analyses: the PRISMA statement report*(16).

### PICO Strategy

P: Aymara Indigenous group and Mapuche Indigenous group. I: Measurement of Body Mass Index and diagnosis of Obesity. C: Comparison between Indigenous and non-indigenous. O: Obesity rate, BMI Mean by group and Odds ratio. Our guiding question was: *Is Aymara or Mapuche indigenous ethnicity a protective/risk factor to obesity?* Our general objective was to identify relationship between indigenous Chilean population and obesity.

### Data analysis

The data obtained were analyzed using meta-analyzes, considering the different populations and established outcomes. Initially, indigenous, and non-indigenous individuals were compared regarding BMI classification. In this analysis, a Random Effect Model was considered, and the Odds Ratio measures obtained using data from the reports. For BMI values, indigenous and non-indigenous individuals were compared, dividing populations into adults and children. Men and women and rural and urban areas were also compared, considering only the indigenous population. In these analyzes, the average differences between the groups were calculated considering a Random Effect Model. In the results of the meta-analyzes, presented by means of forest graphs (forest plot), weighted effect estimates were obtained, as well as their respective 95% confidence intervals and p-values. To assess the presence of heterogeneity, the Chi-square test and the I^2^ statistic were calculated. For the I^2^ statistic, the classification proposed by Higgins and Green(17) was used: -0% to 40%: may not be important; -30% to 60%: may represent moderate heterogeneity; -50% to 90%: can represent substantial heterogeneity; -75% to 100%: considerable heterogeneity. Data were analyzed using Cochrame RevMan 5.3 Software.

## Results

### Systematic Review results

Figure 1 is a schematic representation of search, inclusion, and exclusion of articles. As we collected all the available reports, we analyzed the Systematic Review by subgroups. Due to the difference of the adults and children criteria to perform the diagnosis of obesity, and to reduce clinical heterogeneity, we divided the data obtained into two parts. The first part (Table 1) is data from children and adolescent (6 to 15 years old) and the second part from adolescents and adults (Table 2). Most of the studies identified as adults using the 15 years old as lower age criteria. We excluded 65% of the articles because repeated on different databases and 30% were excluded by Inclusion/Exclusion criteria (Table 3, Fig 1). The remaining 19 articles (Table 1, 2) describe BMI of 1,764,024 individuals.

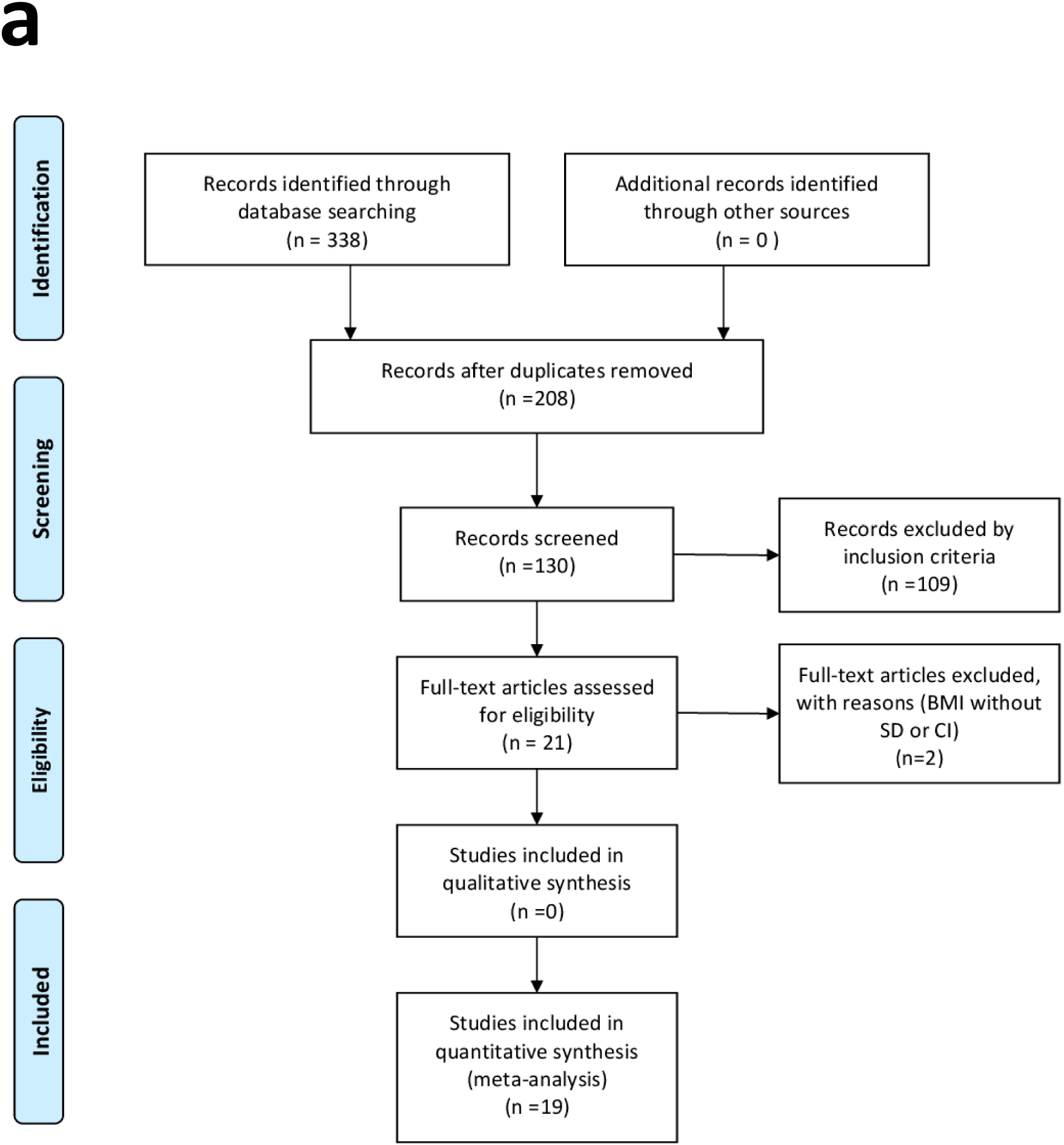

### Meta-analysis results

To explore the relationship between ethnicity and BMI, we compared BMI of Indigenous and non-indigenous. We identified that Indigenous Children showed a lower BMI than Non-Indigenous Children (Fig. 2a). However, we found no difference within BMI adults (Supplementary Fig. S1a).

Also, we explored the relationship between living areas and BMI. We compared the BMI of Indigenous people living in Rural areas and Indigenous people living in Urban areas. We identified that Indigenous people living in rural areas showed lower BMI than those living in urban regions (Fig. 2b)

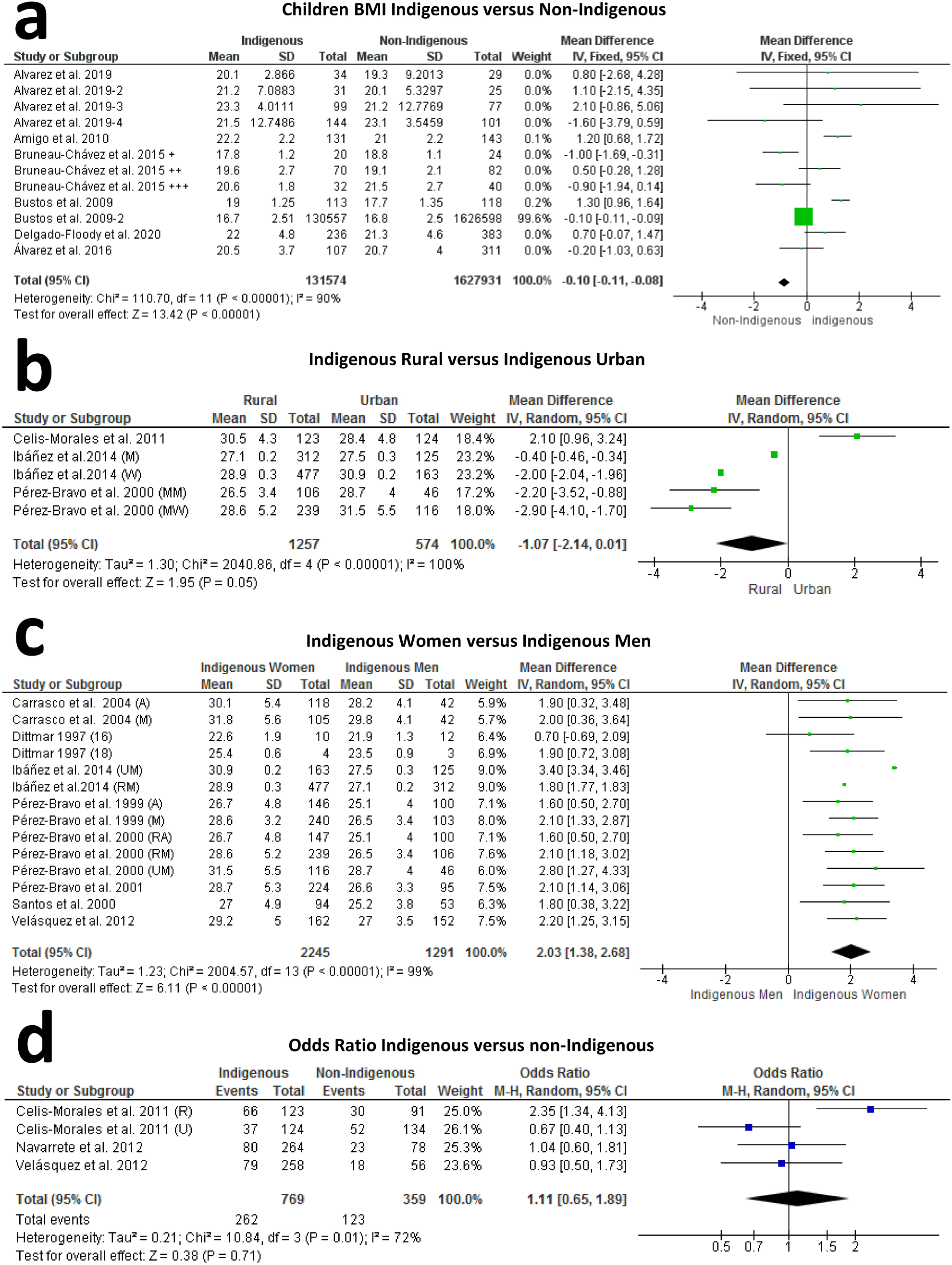

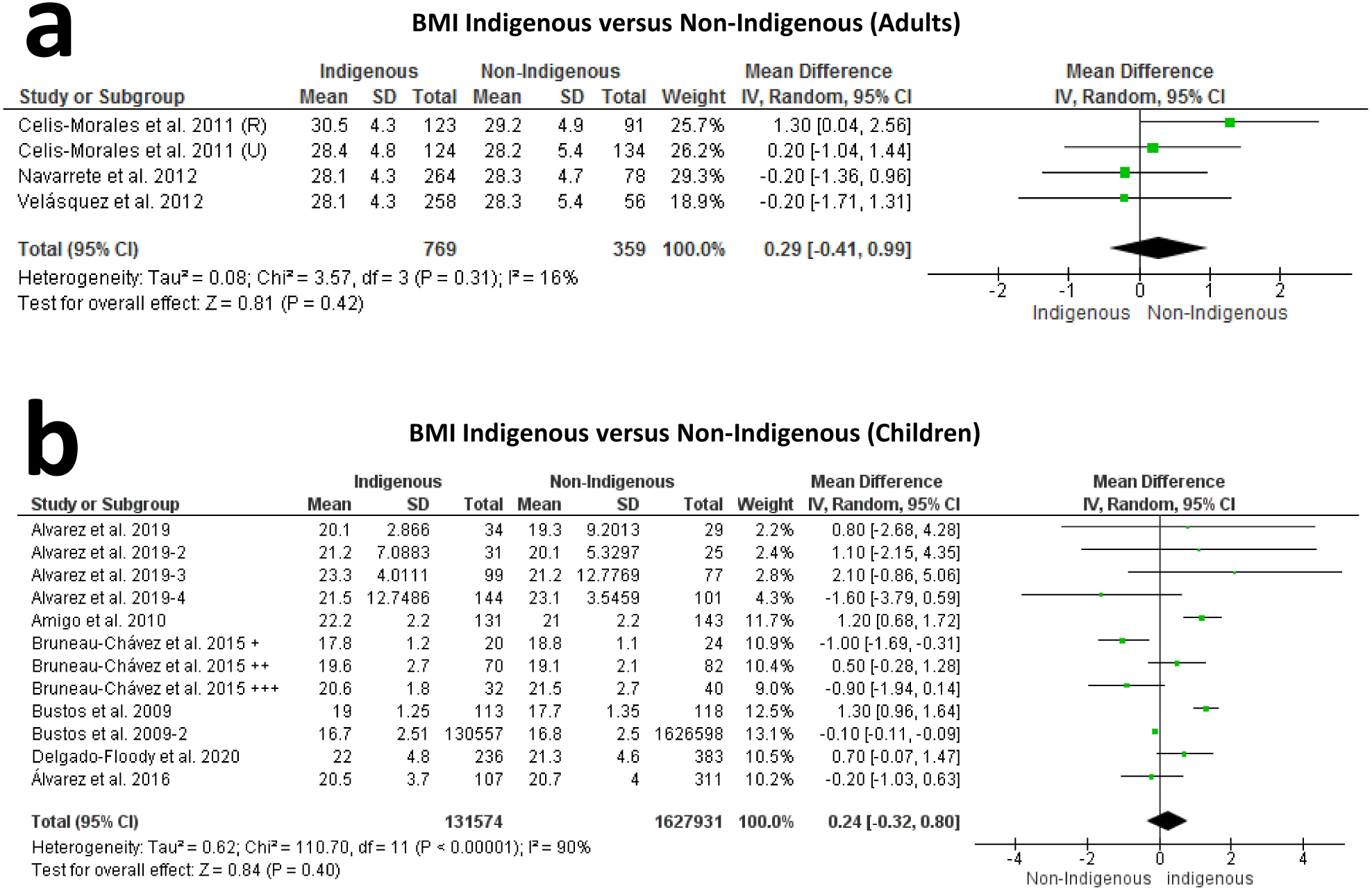

To explore the relationship between gender and BMI, we compared BMI between men and women among indigenous people. We identified that Aymara and Mapuche Women presented higher BMI than Indigenous Men (Fig. 2c).

Finally, to investigate whether ethnicity could represent a risk factor for obesity, we performed am Odds Ratio analysis. We used positive case of obesity among adult groups. We identified no differences in obesity Odds Ratio between Indigenous and non-Indigenous communities (Fig. 2.d).

## Discussion

A constant positive energy balance unavoidably leads to increase BMI and finally to develop obesity. In particular, a positive energy balance is the results from a disparity between energy expenditure and caloric intake (18). This energy conservation equilibrium is conditioned at least by both, the environment (19) and genetic factors (20). Interestingly, people within ancestral ethnic groups share worldview, environmental and genetic features (21) influencing lifestyle and health beliefs (22). Over centuries, most of the indigenous groups worldwide have changed their dietary patterns and lifestyle, once new foods have been introduced or integrated into the traditional food systems by settlers and immigrants. For many years, indigenous groups have lost their access to natural resources and to traditional foods, and have been dislocated from their original place (23). Together, these changes led to a rapid change in the way that they obtained food, facilitating the adoption of unhealthy eating habits (24). As a result, nowadays those native people have been showing the same dietary related-health problems as non-indigenous people, including overweight, obesity, comorbidities, and eating disorders (25).

In some countries, as observed in Australia, indigenous people have social and health disadvantages compared to non-indigenous, what it is important to be considered in the epidemiologic reports on their BMI and eating-related behaviors (27). Independent of body weight gain, the westernization of indigenous health includes the higher consumption of calorically dense foods, increased sodium intake, and lowered nutritional intake of fiber. A study conducted in Panama, observed that 85% of Kuna Indian adults reported eating fast food at least one time weekly, 47% reported eating fried food daily, and 11% reported eating junk food daily (28). With this information, the dietary pattern changes in the last decades should be evaluated in Chilean indigenous as well, throughout validated questionnaires in enough size of a sample.

Beyond the availability of western industrialized foods, the power of choice and of purchasing must be considered in the eating pattern studies with indigenous people. Among Kuna Indians from Panama, for example, those with low-moderate income levels eat junk food and sodas less often; however, they also consume fish and fruits less frequently than those categorized as poor (28). The power of the food manufacturing and social media seems to be extremely relevant for the dietary choices as well.

When thinking about nutrition, it is crucial to consider the meaning of food for each person or group. For indigenous people, the traditional cultivation practices of their food were mostly developed based on continuous observation of the natural life cycle of plants and their connection with the respective environment and ecology (24). It means that they have a spiritual connection with the food, which has also been broken along history. Besides, the healthy eating habit does not include only the food quality but also external factors, like availability for cooking or eating, knowledge for doing healthy choices, and each of this factors is influenced by environmental and cultural changes observed throughout globalization, economic growth, and urbanization, which have been resulting in a more westernized lifestyle (26). In this review, we found that specifically in Mapuche and Aymara, Chilean indigenous groups, there are differences in the risk of obesity development according to gender, age, and social conditions. Indigenous children have a lower BMI than non-indigenous children, as well as rural adult indigenous compared to urban ones. On the other hand, indigenous women have higher BMI when compared to indigenous men. One of the included studies from children showed a larger sample size (Bustos et Munoz, 2009), so a meta-analysis was applied considering all studies in a fixed effect model (Fig. 2a), and another meta-analysis considering a random effect model (Supplementary Fig. 1b).

The Chilean population has three main genetic ancestry contributions: Indigenous (44%), European (52%) and African (4%) (14). Mapuche indigenous group represent the vast majority of all the Chilean Indigenous with 87% of the total indigenous population followed by Aymara indigenous community (4). Aymara and Mapuche surnames have higher Indigenous genetic load, so to determine genetic ancestry in Chilean population, surnames are extremely effective (14). For example, individuals with two Aymara surnames ensure 99.7% of Indigenous polymorphisms contribution, and 0.3% load of European contribution (14). For this reason, we used surname validation to separate indigenous from non-indigenous data.

Previous reports showed Mapuche children presented lower incidence of insulin-dependent diabetes mellitus when compared with non-Indigenous (29). In fact, the presence of Indigenous surname is suggested as a protective factor for T2DM. On the other hand, Aymara rural population that migrated to the urban areas, increased the prevalence of T2DM from 2 to 7% and Rural Mapuche from 4% to 8% (30).

Remains pending for future studies to investigate the relationship between Mapuche and Aymara obesity and Cardiovascular Diseases, as well as the effect of urbanization of Chilean Indigenous groups and Cardiovascular Diseases.

## Limitations

Meta-analyzes presented high statistical heterogeneity and evidence level IV, and then to avoid misleading recommendations new longitudinal research could be required.

## Data Availability

All data is free and available in table 1 and 2. Also, the data were obtained from public available articles.

## Acknowledgements

CPJ was supported by Coordination for the Improvement of Higher Education Personnel (CAPES) grant: 1744875 and 88882.434715/2019-01. NFM was supported by The São Paulo Research Foundation (grant: 2016/17810-3), and LAV and EPA are supported by grants from São Paulo Research Foundation (grants: 2013/07607-8 and 2020/) and Brazilian National Council of Scientific and Technological Development (CNPq).

## Ethics

The study does not require ethical approval because the systematic review is based on published research and the original data are anonymous.

## Bibliography

1. Health Effects of Overweight and Obesity in 195 Countries over 25 Years. New England Journal of Medicine. 2017;377(1):13–27.

2. WHO. Overweight and obesity 2016 [Available from: http://www.who.int/gho/ncd/risk_factors/overweight_text/en/.

3. Trends in adult body-mass index in 200 countries from 1975 to 2014: a pooled analysis of 1698 population-based measurement studies with 19.2 million participants. Lancet. 2016;387(10026):1377–96.

4. Huang T, Shu Y, Cai Y-D. Genetic differences among ethnic groups. BMC genomics. 2015;16:1093-.

5. Tax MG, van der Schoot CE, van Doorn R, Douglas-Berger L, van Rhenen DJ, Maaskant-vanWijk PA. RHC and RHc genotyping in different ethnic groups. Transfusion. 2002;42(5):634–44.

6. Arnaiz-Villena A, Juarez I, Lopez-Nares A, Palacio-Grüber J, Vaquero C, Callado A, et al. Frequencies and significance of HLA genes in Amerindians from Chile Cañete Mapuche. Hum Immunol. 2019;80(7):419–20.

7. Lorenzo Bermejo J, Boekstegers F, González Silos R, Marcelain K, Baez Benavides P, Barahona Ponce C, et al. Subtypes of Native American ancestry and leading causes of death: Mapuche ancestry-specific associations with gallbladder cancer risk in Chile. PLOS Genetics. 2017;13(5):e1006756.

8. Vidal EA, Moyano TC, Bustos BI, Pérez-Palma E, Moraga C, Riveras E, et al. Whole Genome Sequence, Variant Discovery and Annotation in Mapuche-Huilliche Native South Americans. Sci Rep. 2019;9(1):2132.

9. Falconer CL, Park MH, Croker H, Kessel AS, Saxena S, Viner RM, et al. Can the relationship between ethnicity and obesity-related behaviours among school-aged children be explained by deprivation? A cross-sectional study. BMJ Open. 2014;4(1):e003949.

10. Batal M, Decelles S. A Scoping Review of Obesity among Indigenous Peoples in Canada. Journal of obesity. 2019;2019:9741090-.

11. Kirby JB, Liang L, Chen H-J, Wang Y. Race, place, and obesity: the complex relationships among community racial/ethnic composition, individual race/ethnicity, and obesity in the United States. American journal of public health. 2012;102(8):1572–8.

12. Oliveira GF, Oliveira TRR, Ikejiri AT, Galvao TF, Silva MT, Pereira MG. Prevalence of Obesity and Overweight in an Indigenous Population in Central Brazil: A Population-Based Cross-Sectional Study. Obesity Facts. 2015;8(5):302–10.

13. Mitchell BD, Stern MP. Recent developments in the epidemiology of diabetes in the Americas. World Health Stat Q. 1992;45(4):347–9.

14. Fuentes M, Pulgar I, Gallo C, Bortolini M-C, Canizales-Quinteros S, Bedoya G, et al. Geografía génica de Chile: Distribución regional de los aportes genéticos americanos, europeos y africanos. Revista médica de Chile. 2014;142:281–9.

15. Merlin T, Weston A, Tooher R. Extending an evidence hierarchy to include topics other than treatment: revising the Australian ‘levels of evidence’. BMC Medical Research Methodology. 2009;9:34-.

16. Moher D, Liberati A, Tetzlaff J, Altman DG. Preferred reporting items for systematic reviews and meta-analyses: the PRISMA statement. PLoS Med. 2009;6(7):e1000097.

17. Shuster J. Review: Cochrane handbook for systematic reviews for interventions, Version 5.1.0, published 3/2011. Julian P.T. Higgins and Sally Green, Editors. Research Synthesis Methods. 2011;2.

18. Mayer J, Thomas DW. Regulation of food intake and obesity. Science. 1967;156(3773):328–37.

19. Hakala P, Rissanen A, Koskenvuo M, Kaprio J, Ronnemaa T. Environmental factors in the development of obesity in identical twins. Int J Obes Relat Metab Disord. 1999;23(7):746–53.

20. Segal NL, Feng R, McGuire SA, Allison DB, Miller S. Genetic and environmental contributions to body mass index: comparative analysis of monozygotic twins, dizygotic twins and same-age unrelated siblings. International Journal Of Obesity. 2008;33:37.

21. The Use of Racial, Ethnic, and Ancestral Categories in Human Genetics Research. The American Journal of Human Genetics. 2005;77(4):519–32.

22. Alarcon AM, Vidal A, Castro M. Cultural meanings of musculoskeletal diseases in Chile’s Mapuche Population. J Transcult Nurs. 2013;24(4):340–7.

23. Clark TD. Putting the market in its place: food security in three Mapuche communities in southern Chile. Latin American research review. 2011;46(2):154–79.

24. Sarkar D, Walker-Swaney J, Shetty K. Food Diversity and Indigenous Food Systems to Combat Diet-Linked Chronic Diseases. Current Developments in Nutrition. 2019;4(Supplement_1):3–11.

25. Herbozo S, Flynn PM, Stevens SD, Betancourt H. Dietary Adherence, Glycemic Control, and Psychological Factors Associated with Binge Eating Among Indigenous and Non-Indigenous Chileans with Type 2 Diabetes. International journal of behavioral medicine. 2015;22(6):792–8.

26. Riesco M. Latin America: a new developmental welfare state model in the making?1. International Journal of Social Welfare. 2009;18(1):S22–S36.

27. Hardy LL, MacNiven R, Esgin T, Mihrshahi S. Cross-sectional changes in weight status and weight related behaviors among Australian children and Australian Indigenous children between 2010 and 2015. PloS one. 2019;14(7):e0211249–e.

28. Neitzel AL, Smalls BL, Walker RJ, Dawson AZ, Campbell JA, Egede LE. Examination of dietary habits among the indigenous Kuna Indians of Panama. Nutrition journal. 2019;18(1):44-.

29. Larenas G, Montecinos A, Manosalva M, Barthou M, Vidal T. Incidence of insulin-dependent diabetes mellitus in the IX region of Chile: ethnic differences. Diabetes Res Clin Pract. 1996;34 Suppl:S147–51.

30. Carrasco EP, Perez FB, Angel BB, Albala CB, Santos JL, Larenas GY, et al. [Prevalence of type 2 diabetes and obesity in two Chilean aboriginal populations living in urban zones]. Rev Med Chil. 2004;132(10):1189–97.

