## Supplementary material for "SOUTH AMERICAN INDIGENOUS COMMUNITIES AND BODY MASS INDEX: A SYSTEMATIC REVIEW AND META-ANALYSIS": Table 3

**Table 1**. Summary of Identification, screening and eligibility process.

| **Database** | **Found** | **Excluded 1*** | **Excluded 2**** | **Final sample** |
| --- | --- | --- | --- | --- |
| LILACS | 79 | 59 | 18 | 3 |
| SciELO | 36 | 27 | 7 | 2 |
| PubMed | 67 | 25 | 32 | 7 |
| Scopus | 80 | 58 | 20 | 2 |
| Web of Science | 76 | 39 | 32 | 5 |
| **Total** | **338** | **208** | **109** | **19** |

*Number of articles excluded by repeated, or ** did not fit to inclusion criteria
