## Supplementary material for "SOUTH AMERICAN INDIGENOUS COMMUNITIES AND BODY MASS INDEX: A SYSTEMATIC REVIEW AND META-ANALYSIS": Table 1 and 2

Summary of descriptive characteristics of included articles (n=16)

**Table 1: Children**

| Source | Publication  Year | No. of Participants | Indigenous and Obesity criteria | BMI: Mean (SD) or (CI) | Age: y mean (CI) or (SD) | Levels of Evidence | Significance |
| --- | --- | --- | --- | --- | --- | --- | --- |
| Bustos et al.  (1) | 2009 | 231 Girls  113 Indigenous  118 Non-Indigenous | Mapuche  2 surnames | Indigenous: 19,0 (17,5-20,0)  Non-Indigenous: 17,7 (16,6-19,3) | 10,4 (9,7-10,9)  10,4 (9,6 – 10,7) | III | 0,001** |
| Bustos et al.  (2) | 2009 | 130.557 Mapuche  1.626.598 Non-Mapuche | Mapuche  2 surnames | Indigenous: 16.7 (2.51)  Non-Indigenous: 16.77 (2.5) | 6,35 (0,36)  6,37 (0,38) | IV | < 0.01 |
| Amigo et al.  (3) | 2010 | 274 Girls  131 Indigenous  143 Non-Indigenous | Mapuche  2 surnames | Indigenous: 22.2 (20.1-24.5)  Non-Indigenous: 21.0 (19.2-23.6) | 12,3 (9–16) | IV | 0,069** |
| Bruneau-Chávez et al.  (4) | 2015 | 268 Boys  20 Indigenous  24 Non-Indigenous  70 Indigenous  82 Non-Indigenous  32 Indigenous  40 Non-Indigenous | Mapuche  2 surnames | Indigenous: 17.8 (1.2)  Non-Indigenous: 18.8 (1.1)  Indigenous: 19.6 (2.7)  Non-Indigenous: 19.1 (2.1)  Indigenous: 20.6 (1.8)  Non-Indigenous: 21.5 (2.7) | 10  11-12  13 | IV | > 0,05  > 0,05  > 0,05 |
| Álvarez et al.  (5) | 2016 | 241 Girls  177 Boys  107 Indigenous  311 Non-Indigenous | Mapuche  ≥1 surname | Indigenous: 20,5 (3,7)  Non-Indigenous: 20,7 (4,0) | 9,5 (2,5)  9,6 (2,4) | IV | 0,629 |
| Valdés-Badilla et al.  (6) | 2015 | 11 Mapuche Girls  12 Mapuche Boys | Mapuche  ≥1 surname | Mapuche Girls: 17,9 (2,6)  Mapuche Boys: 19,5 (3,7) | 8,9 (1,6)  8,4 (1,4) | IV | - |
| Delgado-Floody et al.  (7) | 2020 | 236 Mapuche  383 Non-Mapuche  104 Mapuche Girls  132 Mapuche Boys  169 Non-Mapuche Girls  214 Non-Mapuche Boys | Mapuche  2 surnames | Indigenous: 22.0 (4.8)  Non-Indigenous: 21.3 (4,6) | 11.64 (1,0)  11.77 (1.1) |  |  |
| Alvarez et al.  (8) | 2019 | 34 Mapuche  29 Non-Mapuche  31 Mapuche  25 Non-Mapuche  99 Mapuche  77 Non-Mapuche  144 Mapuche  101 Non-Mapuche | Mapuche  2 surnames | 20.1 (18.9 - 21.1)  19.3 (18.2 – 22.8)  21.2 (19.2 – 23.8)  20.1 (18.6 – 22.3)  23.3 (22.5 - 24.1)  21.2 (19.1 – 24.1)  21.5 (19.0 – 23.6)  23.1 (22.7 – 23.8) | 9.7 (7.9-11.5)  11.8 (10.3-13.2)  10.0 (7.4-12.5)  10.8 (9.5-12.1)  8.6 (6.6 – 10.7)  8.9 (8.2 - 9.6)  11.5 (9.7-13.2)  9.1 (8.2-10.0) |  |  |
| Dittmar  (9) | 1997 | 9 Aymara Women  6 Aymara Men  17 Aymara Women  5 Aymara Men  10 Aymara Women  10 Aymara Men  15 Aymara Women  20 Aymara Men  11 Aymara Women  21 Aymara Men | Aymara  2 surnames | Aymara Women: 15.5 (1.1)  Aymara Men: 16.6 (1.9)  Aymara Women: 16.4 (1.4)  Aymara Men: 16.5 (0.64)  Aymara Women: 17.9 (1.0)  Aymara Men: 17.2 (0.9)  Aymara Women: 19.8 (2.2)  Aymara Men: 19.6 (1.6)  Aymara Women: 21.0 (1.9)  Aymara Men: 20.4 (1.85) | 6.0-7.9  8.0-9.9  10.0-11.9  12.0-13.9  14.0-15.9 | IV | - |

**Table 2: Adults**

| Dittmar  (9) | 1997 | 10 Aymara Women  12 Aymara Men  4 Aymara Women  3 Aymara Men | Aymara  2 surnames | Aymara Women: 22.6 (1.9)  Aymara Men: 21.9 (1.3)  Aymara Women: 25.4 (0.6)  Aymara Men: 23.5 (0.9) | 16.0-17.9  18.0-19.9 | IV | - |
| --- | --- | --- | --- | --- | --- | --- | --- |
| Velásquez et al.  (10) | 2012 | 131 Indigenous Women  127 Indigenous Men  31 Non-Indigenous Women  25 Non-Indigenous Men  258 Indigenous  56 Non-Indigenous | Pehuenche  2 surnames*  BMI ≥ 30 | Indigenous: 28,1 (4,3)  Non-Indigenous: 28,3 (5,4)  Pehuenche Women: 29.2 (5.0)  Pehuenche Men: 27.0 (3.5) | > 18 | IV | 0,77 |
| Navarrete et al.  (11) | 2012 | 134 Indigenous Women  130 Indigenous Men  46 Non-Indigenous Women  32 Non-Indigenous Men  264 Indigenous  78 Non-Indigenous | Pehuenche  2 surnames*  BMI ≥ 30 | Indigenous: 28,1 (4,3)  Non-Indigenous: 28,3 (4,7) | 42,2 (15,1)  37,1 (14) | III | 0,970 |
| Celis-Morales et al.  (12) | 2011 | 69 Rural Mapuche Women  54 Rural Mapuche Men  79 Urban Mapuche Women  45 Urban Mapuche Men  55 Rural Non-Mapuche Women  36 Rural Non-Mapuche Men  92 Urban Non-Mapuche Women  42 Urban Non-Mapuche Men | Mapuche  2 surnames  BMI ≥ 30 | Rural Mapuche: 30.5 (4.3)  Urban Mapuche: 28.4 (4.8)  Rural Non-Mapuche: 29.2 (4.9)  Urban Non-Mapuche: 28.2 (5.4) | 36,7 (11,9)  34,1 (12,5)  40,9 (13,7)  37,5 (12,4) | IV | < 0,05 |
| Ibáñez et al.  (13) | 2014 | 640 Women  437 Men  477 Rural Mapuche Women  312 Rural Mapuche Men  163 Urban Mapuche Women  125 Urban Mapuche Men | Mapuche  2 surnames*  Central Obesity | Rural Mapuche Women: 28.9 (0.3)  Rural Mapuche Men: 27.1 (0.2)  Urban Mapuche Women: 30.9 (0.2)  Urban Mapuche Men: 27.5 (0.3) | 43,1 (0,6)  43,8 (0,7)  42,6 (1,1)  37,1 (1,3) | IV | < 0,05 |
| Pérez-Bravo et al.  (14) | 2001 | 224 Mapuche Women  95 Mapuche Men | Mapuche  2 surnames  BMI ≥ 25 | Mapuche Women: 28,7 (5.3)  Mapuche Men: 26.6 (3.3) | > 20 | IV | 0.005 |
| Pérez-Bravo et al.  (15) | 2000 | 239 Rural Mapuche Women  106 Rural Mapuche Men  147 Rural Aymara Women  100 Rural Aymara Men  116 Urban Mapuche Women  46 Urban Mapuche Men | Mapuche and Aymara  2 surnames  BMI ≤ 30  (HANNES Index) | Rural Mapuche Women: 28,6 (5,2)  Rural Mapuche Men: 26,5 (3,4)  Rural Aymara Women: 26,7 (4,8)  Rural Aymara Men: 25,1 (4,0)  Urban Mapuche Women: 31,5 (5,5)  Urban Mapuche Men: 28.7 (4,0) | 47,5 (16,1)  48,5 (15,2)  49,1 (17,7)  48,4 (19,2)  44,5 (14,9)  48,5 (14,6) | IV | < 0,05 |
| Pérez-Bravo et al.  (16) | 1999 | 240 Mapuche Women  103 Mapuche Men  146 Aymara Women  100 Aymara Men | Mapuche and Aymara  2 surnames  BMI > 30 | Mapuche Women: 28,6 (3,2)  Mapuche Men: 26,5 (3,4)  Aymara Women: 26,7 (4,8)  Aymara Men: 25,1 (4,0) | 47,5 (16,1)  48,5 (15,2)  45,1 (17,7)  48,4 (19,2) | IV | < 0,05 |
| Carrasco et al.  (17) | 2004 | 105 Urban Mapuche Women  42 Urban Mapuche Men  118 Urban Aymara Women  42 Urban Aymara Men | Mapuche and Aymara  2 surnames  BMI ≥ 30 | Mapuche Women: 31,8 (5,6)  Mapuche Men: 29,8 (4,1)  Aymara Women: 30,1 (5,4)  Aymara Men: 28,2 (4,1) | 44,3 (14,2)  45,6 (13,9)  47,5 (13,6)  52,1 (14,1) | IV | p <0,04 |
| Santos et al.  (18) | 2000 | 94 Aymara Women  53 Aymara Men | Aymara  2 surnames  BMI ≥ 25 | Aymara Women: 27,0 (4,9)  Aymara Men: 25,2 (3,8) | 45,0 (17,5)  49,4 (18,3) | IV | 0.02 |

Age in years. * Validated by the National Indigenous Development Corporation of Chile **Median Test. BMI: kg/m^2^. Level of evidence based in the *NHMRC Evidence Hierarchy*

1.	Bustos P, Amigo H, Muzzo S, Ossa X. [Thelarche and nutritional status: an epidemiological study of two ethnic groups]. Rev Med Chil. 2009;137(10):1301-8.

2.	Bustos P, Munoz S, Vargas C, Amigo H. Evolution of the nutritional situation of indigenous and non-indigenous Chilean schoolchildren. Ann Hum Biol. 2009;36(3):298-307.

3.	Amigo H, Bustos P, Muzzo S, Alarcon AM, Munoz S. Age of menarche and nutritional status of indigenous and non-indigenous adolescents in the Araucania Region of Chile. Ann Hum Biol. 2010;37(4):554-61.

4.	Bruneau-Chávez J, España-Romero V, Lang-Tapia M, Chillón Garzón P. Diferencias en la Composición Corporal y Somatotipo de Escolares de Etnia Mapuche y no Mapuche de la Comuna de Temuco - Chile. International Journal of Morphology. 2015;33:988-95.

5.	Álvarez C, Ramírez-Campillo R, Martínez-Salazar C, Vallejos-Rojas A, Jaramillo-Gallardo J, Salas Bravo C, et al. Hipertensión en relación con estado nutricional, actividad física y etnicidad en niños chilenos entre 6 y 13 años de edad. Nutrición Hospitalaria. 2016;33:220-5.

6.	Valdés-Badilla PA, Vergara-Coronado NY, Suazo-Poblete D, Godoy-Cumillaf A, Herrera-Valenzuela T, Durán-Agüero S. Perfil antropométrico y hábitos de actividad física de estudiantes Mapuches de una escuela rural de Temuco, Chile. Revista Española de Nutrición Humana y Dietética. 2015;19:28-35.

7.	Delgado-Floody P, Caamaño-Navarrete F, Guzmán-Guzmán IP, Jerez-Mayorga D, Martínez-Salazar C, Álvarez C. Food Habits and Screen Time Play a Major Role in the Low Health Related to Quality of Life of Ethnic Ascendant Schoolchildren. Nutrients. 2020;12(11).

8.	Álvarez C, Lucia A, Ramírez-Campillo R, Martínez-Salazar C, Delgado-Floody P, Cadore EL, et al. Low sleep time is associated with higher levels of blood pressure and fat mass in Amerindian schoolchildren. Am J Hum Biol. 2019;31(6):e23303.

9.	Dittmar M. Linear growth in weight, stature, sitting height and leg length, and body proportions of Aymara school-children living in an hypoxic environment at high altitude in Chile. Z Morphol Anthropol. 1997;81(3):333-44.

10.	Cartes Velásquez R, Navarrete Briones C. Caracterización antropométrica de población pehuenche adulta, consideraciones nutricionales. Alto Biobio, Chile. Memorias del Instituto de Investigaciones en Ciencias de la Salud. 2012;10:30-7.

11.	Navarrete B C, Cartes-Velásquez R. Prevalencia de diabetes tipo 2 y obesidad en comunidades Pehuenches, Alto Biobio. Revista chilena de nutrición. 2012;39:7-10.

12.	Celis-Morales CA, Perez-Bravo F, Ibanes L, Sanzana R, Hormazabal E, Ulloa N, et al. Insulin resistance in Chileans of European and indigenous descent: evidence for an ethnicity x environment interaction. PLoS One. 2011;6(9):e24690.

13.	Ibáñez L, Sanzana R, Salas C, Navarrete C, Cartes-Velásquez R, Rainqueo A, et al. Prevalencia de síndrome metabólico en individuos de etnia Mapuche residentes en zonas rurales y urbanas de Chile. Revista médica de Chile. 2014;142:953-60.

14.	Perez-Bravo F, Carrasco E, Santos JL, Calvillan M, Larenas G, Albala C. Prevalence of type 2 diabetes and obesity in rural Mapuche population from Chile. Nutrition. 2001;17(3):236-8.

15.	Pérez B F, Santos M JL, Albala B C, Calvillán C M, Carrasco P E. Asociación obesidad y leptina en tres poblaciones aborígenes de Chile. Revista médica de Chile. 2000;128:45-52.

16.	Pérez B F, Carrasco P E, Santos JL, Calvillán M, Albala B C. Prevalencia de obesidad , hipertensión arterial y dislipidemia en grupos aborígenes rurales de Chile. Revista médica de Chile. 1999;127:1169-75.

17.	Carrasco EP, Perez FB, Angel BB, Albala CB, Santos JL, Larenas GY, et al. [Prevalence of type 2 diabetes and obesity in two Chilean aboriginal populations living in urban zones]. Rev Med Chil. 2004;132(10):1189-97.

18.	Santos JL, Perez-Bravo F, Albala C, Calvillan M, Carrasco E. Plasma leptin and insulin levels in Aymara natives from Chile. Ann Hum Biol. 2000;27(3):271-9.
